# Exceptionally low likelihood of Alzheimer’s dementia in APOE2 homozygotes

**DOI:** 10.1101/19011015

**Authors:** Eric M. Reiman, Joseph F. Arboleda-Velasquez, Yakeel T. Quiroz, Matthew J. Huentelman, Thomas G. Beach, Richard J. Caselli, Yinghua Chen, Yi Su, Amanda J. Myers, John Hardy, Jean Paul Vonsattel, Steven G. Younkin, David A. Bennett, Philip L. De Jager, Eric B. Larson, Paul K. Crane, C. Dirk Keene, M. Ilyas Kamboh, Julia K. Kofler, Linda Duque, John R. Gilbert, Harry E. Gwirtsman, Joseph D. Buxbaum, Dennis W. Dickson, Matthew P. Frosch, Bernardino Ghetti, Kathryn L. Lunetta, Li-San Wang, Bradley T. Hyman, Walter A. Kukull, Tatiana Foroud, Jonathan L. Haines, Richard P. Mayeux, Margaret A. Pericak-Vance, Julie A. Schneider, John Q. Trojanowski, Lindsay A. Farrer, Gerard D. Schellenberg, Gary W. Beecham, Thomas J. Montine, Gyungah R. Jun, for the Alzheimer’s Disease Genetics Consortium

## Abstract

Each additional copy of the apolipoprotein E4 (APOE4) allele is associated with a higher risk of Alzheimer’s dementia, such that APOE4 homozygotes have a particularly high risk. While the APOE2 allele is associated with a lower risk of Alzheimer’s dementia, it is not yet known whether APOE2 homozygotes have a particularly low risk. We generated Alzheimer’s dementia odds ratios and other findings in more than 5,000 clinically characterized and neuropathologically characterized Alzheimer’s dementia cases and controls. APOE2/2 was associated with exceptionally low Alzheimer’s dementia odds ratios compared to APOE2/3, 3/3 and 4/4, and the impact of APOE2 and APOE4 gene dose was significantly greater in the neuropathologically confirmed group than in more than 24,000 neuropathologically unconfirmed cases and controls. Finding and targeting the factors by which APOE and its variants influence Alzheimer’s disease could have a major impact on the understanding, treatment and prevention of this terrible disease.

## INTRODUCTION

Apolipoprotein E (*APOE*), the major susceptibility gene for late-onset Alzheimer’s disease (AD), has three common alleles (APOE2, 3, and 4), giving rise to six genotypes (APOE2/2, 2/3, 3/3, 2/4, 3/4, and 4/4). Compared to the most common APOE3/3 genotype, each additional copy of the APOE4 allele is associated with a higher risk of Alzheimer’s dementia and a younger mean age at dementia onset, such that APOE4 homozygotes are at the highest risk, while the presence of one or two copies of the APOE2 allele is associated with a lower risk of Alzheimer’s dementia and an older mean age at dementia onset.^1-4^ It remains to be clarified whether APOE2 homozygotes have a lower odd than persons with the APOE2/3 genotype—a question we sought to address in an unusually large number of clinically and neuropathologically classified Alzheimer’s dementia cases and controls.

We (JFA-V, YQ, GRJ, EMR), Francisco Lopera, and our colleagues recently discovered two copies of the rare APOE3 Christchurch (APOE3ch [Arg136→Ser]) mutation in an amyloid-β_42_ (Aβ_42_) overproducing presenilin 1 (PSEN1) E280A mutation carrier from the world’s largest autosomal dominant AD kindred who did not develop mild cognitive impairment (MCI) until her seventies, nearly three decades after her kindred’s mean age at MCI onset.^5^ Using PET to compare her to other PSEN1 E280A mutation carriers with MCI, she had the greatest fibrillar amyloid-β (Aβ) burden (the major constituent of neuritic plaques), limited paired helical filament tau (neurofibrillary tangle) burden, and minimal glucose hypometabolism in brain regions preferentially affected by AD. Like 5-10% of APOE2 homozygotes, she also had hyperlipoproteinemia Type III, reflecting reduced ApoE protein low density lipoprotein receptor (LDLR) binding.5,6 Like the ApoE2 protein, the ApoEch protein was associated with less Aβ_42_ aggregation than the ApoE3 protein *in vitro*.^5,7^ These findings led us to postulate that APOE2 homozygotes might have an exceptionally low risk of late-onset Alzheimer’s dementia.

Why has this possibility remained unaddressed? First, case-control studies without neuropathological or biomarker assessments of AD may have underestimated the impact of APOE genotypes on Alzheimer’s dementia odds ratios (ORs) due to the confounding effects of APOE genotypes on the percentages of neuropathologically misclassified cases and controls.^8,9,10^ Second, previous studies of clinically and neuropathologically characterized cases and controls may have been too small to demonstrate that APOE2 homozygotes have an even lower OR than the relatively low-risk APOE2/3 group due to the paucity of APOE2 homozygotes, who comprise less than 1% of the general population.^1,4^

This study sought to establish that APOE2 homozygotes have an exceptionally low likelihood of Alzheimer’s dementia, demonstrate the value of AD risk assessments in clinically and neuropathologically characterized cases and controls, and underscore the impact of different *APOE* genotypes on Alzheimer’s dementia ORs relative to the lowest risk APOE2/2 and highest risk APOE4/4 genotypes. More generally, it sought to highlight the impact of discovering and targeting the mechanism by which *APOE* variants account for differential risk could have on the understanding, treatment and prevention of AD, including those interventions that might prevent both the initial development of AD pathology and the subsequent development of dementia.

## RESULTS

### Neuropathologically confirmed, unconfirmed, and combined groups

***Supplementary Table 1*** shows the number of Alzheimer’s dementia cases and cognitively unimpaired controls for each APOE genotype in a) the ADGC’s clinically characterized and neuropathologically confirmed autopsy group, b) its clinically characterized but neuropathologically unconfirmed non-autopsy group, and c) the combined neuropathologically confirmed and unconfirmed group. The 5,007 participants in the neuropathologically confirmed cohort included 4,018 AD dementia cases and 989 cognitively unimpaired and neuropathologically unaffected controls. The 23,857 participants in the clinically classified but neuropathologically unconfirmed cohort included 10,430 probable AD dementia cases and 13,426 cognitively unimpaired controls. The 28,864 participants in the combined group included 14,448 cases and 14,416 controls. ***Supplementary Table 2*** summarizes ages at dementia onset in the cases, ages at last clinical exam in the cases, and ages at death in the neuropathologically confirmed autopsy cohort.

### Alzheimer’s Dementia ORs

***Table 1 and Supplementary Table 3*** show Alzheimer’s dementia ORs for each APOE genotype and allelic doses (i.e., the number of APOE2 alleles in APOE4 non-carriers and number of APOE4 alleles in APOE2 non-carriers) before and after adjustment for age and sex in the neuropathologically confirmed and unconfirmed groups before and after adjustment for age and sex, compared to the common APOE3/3 genotype. As discussed below, APOE2/2, APOE2/3, and APOE2 allelic dose ORs were significantly lower, and APOE3/4, APOE4/4, and APOE4 allelic dose ORs were significantly higher, in the neuropathologically confirmed than unconfirmed group. ***Table 2*** shows Alzheimer’s dementia ORs for each APOE genotype compared to the relatively low risk APOE2/3 and highest risk APOE4 genotypes in the neuropathologically confirmed cohort. As discussed below, these ORs permitted us to confirm our primary hypothesis that APOE2/2 is associated with a significantly lower OR compared to APOE2/2 and to demonstrate an exceptionally low OR compared to APOE4/4. ***Supplementary Table 4*** shows Alzheimer’s dementia ORs for each APOE genotype in the combined group, compared to APOE3/3, and for APOE2 and APOE4 allelic dose before and after adjustment for age, sex, and autopsy/non-autopsy group.

**Table 1.** Alzheimer’s Dementia Odds Ratios for Each APOE genotype, Compared to APOE3/3, and for APOE2 and APOE4 Allelic Doses in the ADGC’s Neuropathologically Confirmed and Unconfirmed Groups. The ADGC’s “neuropathologically confirmed group” includes cases who met clinical and neuropathological criteria for Alzheimer’s dementia and controls in the who were cognitively unimpaired at their last clinical exam and did not meet neuropathological criteria for AD at autopsy. The ADGC’s neuropathologically unconfirmed group includes cases who met clinical criteria for Alzheimer’s dementia and cognitively unimpaired controls, who were included irrespective of any antemortem biomarker or subsequent post-mortem neuropathological assessments they may have had. OR refers to odds ratio. 95% CI refers to 95% confidence interval ORs for each APOE genotype were calculated using genotypic association tests compared to the APOE3/3 genotype as the reference in an additive genetic model. ORs associated with APOE2 allelic dose in APOE4 non-carriers (APOE2/2<2/3<3/3) and APOE4 allelic dose in APOE2 non-carriers (APOE4/4>3/4>3/3) were generated using allelic association tests in an additive genetic model. The number and percentage of cases and controls, ages at estimated clinical onset or last clinical exam, and ages at death are shown for each APOE genotype, are shown for each group in Supplementary Tables 1 and 2.

| *APOE* | Neuropath Confirmed Group |  |  | Neuropath Unconfirmed Group |  |  |
| --- | --- | --- | --- | --- | --- | --- |
|  | OR | 95% CI | P | OR | 95% CI | P |
| **Genotype** |  |  |  |  |  |  |
| **2/2** | 0.13 | 0.05 - 0.36 | $6.3 \times 10^{-5}$ | 0.52 | 0.30 - 0.90 | 0.02 |
| **2/3** | 0.39 | 0.30 - 0.50 | $1.6 \times 10^{-12}$ | 0.63 | 0.53 - 0.75 | $2.2 \times 10^{-7}$ |
| **2/4** | 2.68 | 1.65 - 4.36 | $7.5 \times 10^{-5}$ | 2.47 | 2.02 - 3.01 | $5.7 \times 10^{-19}$ |
| **3/4** | 6.13 | 5.08 - 7.41 | $2.2 \times 10^{-75}$ | 3.55 | 3.17 - 3.98 | $2.3 \times 10^{-105}$ |
| **4/4** | 31.22 | 16.59 - 58.75 | $4.9 \times 10^{-26}$ | 10.70 | 9.12 - 12.56 | $7.5 \times 10^{-186}$ |
| **Allelic Dose** |  |  |  |  |  |  |
| **2** | 0.38 | 0.30 - 0.48 | $1.1 \times 10^{-15}$ | 0.64 | 0.58 - 0.72 | $2.2 \times 10^{-16}$ |
| **4** | 6.00 | 5.06 - 7.12 | $3.4 \times 10^{-90}$ | 3.43 | 3.26 - 3.60 | $< 10^{-300}$ |

**Table 2.** Alzheimer’s Dementia Odds Ratios for each APOE Genotype, Compared to APOE2/3 and 4/4, in the Neuropathologically Confirmed Group.

| ***APOE*** | **Compared to APOE2/3** |  |  | **Compared to APOE4/4** |  |  |
| --- | --- | --- | --- | --- | --- | --- |
|  | **OR** | **95% CI** | **P** | **OR** | **95% CI** | **P** |
| **2/2** | 0.34 | 0.12 - 0.95 | 0.04 | 0.004 | 0.001 - 0.014 | 6.0 x10<sup>-19</sup> |
| **2/3** | 1.00 | - | - | 0.012 | 0.006 - 0.024 | 1.2 x10<sup>-34</sup> |
| **3/3** | 2.60 | 2.00 - 3.38 | 1.6 x10<sup>-12</sup> | 0.032 | 0.017 - 0.060 | 4.9 x10<sup>-26</sup> |
| **2/4** | 6.96 | 4.06 - 11.92 | 7.5 x10<sup>-12</sup> | 0.086 | 0.039 - 0.189 | 1.6 x10<sup>-9</sup> |
| **3/4** | 15.92 | 11.85 - 21.38 | 1.4 x10<sup>-70</sup> | 0.196 | 0.103 - 0.375 | 8.4 x10<sup>-7</sup> |
| **4/4** | 81.05 | 41.39 - 158.68 | 1.2 x10<sup>-34</sup> | 1.000 | - | - |

**Table 3.** Residual Effects of Each APOE Genotype on Braak (Tau Tangle) Stage, after Adjustment for CERAD (Neuritic Aβ Plaque) Score.

| **APOE** | **Compared to APOE2/3** |  |  | **Compared to APOE3/3** |  |  | **Compared to APOE4/4** |  |  |
| --- | --- | --- | --- | --- | --- | --- | --- | --- | --- |
|  | **BETA\*** | **SE** | **P** | **BETA\*** | **SE** | **P** | **BETA\*** | **SE** | **P** |
| **2/2** | -0.44 | 0.31 | 0.15 | -0.50 | 0.28 | 0.07 | -1.32 | 0.27 | 1.6 x10<sup>-6</sup> |
| **2/3** | - | - | - | -0.07 | 0.09 | 0.44 | -0.43 | 0.11 | 1.2 x10<sup>-4</sup> |
| **3/3** | 0.07 | 0.09 | 0.39 | - | - | - | -0.25 | 0.06 | 4.3 x10<sup>-5</sup> |
| **2/4** | 0.13 | 0.16 | 0.40 | 0.03 | 0.12 | 0.80 | -0.26 | 0.11 | 0.02 |
| **3/4** | 0.29 | 0.08 | 6.1 x10<sup>-4</sup> | 0.15 | 0.04 | 1.9 x10<sup>-4</sup> | -0.10 | 0.05 | 0.05 |
| **4/4** | 0.42 | 0.10 | 7.5 x10<sup>-5</sup> | 0.25 | 0.06 | 4.3 x10<sup>-5</sup> | - | - | - |

\* Beta estimates in a linear regression model, relative to APOE2/3, 3/3, and 4/4. Beta coefficients reflect the impact of each genotype on Braak stage after adjustment for CERAD score. Positive and negative beta coefficients correspond to increased or decreased Braak stage, respectively. Findings do not account for potentially confounding effects of age at death, clinical duration prior to death, or sex.

#### Impact of APOE2 homozygosity

As shown in ***Tables 1 and 2***, APOE2 homozygotes had a significantly lower 0.34 OR (95% CI=0.12-0.95) compared to the relatively low-risk APOE2/3 genotype, an extremely low 0.13 OR (95% CI 0.05-0.36) compared to the most common APOE3/3 genotype; and an exceptionally low 0.004 OR (95% CI 0.001-0.014) compared to the highest risk APOE4/4 genotype. APOE2 homozygotes had more modest but significantly lower ORs compared to APOE3/3 in both the clinical group (0.52 OR [95% CI=0.30-0.90] and (not shown) in the combined group (0.41 OR [95% CI=0.25-0.66], P=3×10^−4^). As shown in ***Supplementary Table 3***, ORs for each APOE genotype, compared to APOE3/3, were not significantly affected in the neuropathologically confirmed or unconfirmed group after adjustment for age and sex.

#### Impact of APOE2 allelic dose

As shown in ***Table 1***, APOE2 allelic dose in APOE4 non-carriers was associated with significantly lower ORs in both the neuropathologically confirmed and unconfirmed groups, and with significantly lower ORs in the confirmed than unconfirmed group (P=6×10^−4^). Using an additive genetic model, ORs in the neuropathologically confirmed (0.38 OR [95% CI=0.30-0.48] P=1×10^−15^), unconfirmed (0.64 OR [95% CI=0.58-0.72] P=2×10^−16^), and combined groups (0.59 OR [95% CI=0.53-0.65] P=9×10^−27^) were similar to the corresponding APOE2/3 ORs relative to APOE3/3. As shown in ***Supplementary Tables 3 and 4***, findings were not significantly affected after adjustment for age, sex, and neuropathologically confirmed autopsy/unconfirmed non-autopsy group. Using the resulting beta estimates, the 0.14 OR in the neuropathologically confirmed APOE2 homozygotes was similar to the 0.13 OR shown in Table 1.

#### Impact of other APOE genotypes

As previously noted, compared to APOE3/3, APOE2/2 and 2/3 genotypes were associated with significantly lower Alzheimer’s dementia ORs, and APOE3/4 and 4/4 genotypes were associated with significantly higher ORs, in the neuropathologically confirmed than unconfirmed or combined groups than in the neuropathologically unconfirmed or combined groups (P<10^−4^). APOE2/4, APOE3/4, and APOE4/4 genotypes in the neuropathologically confirmed autopsy group were associated with 2.68, 6.13, and 31.22 ORs, respectively, compared to the most common APOE3/3 genotype; 20.33, 46.51, and 236.74, respectively, compared to the lowest risk APOE2/2 genotype; and 0.09, 0.20, and 1.00, respectively, compared to the highest odds APOE4/4 genotype. These results underscore the impact of *APOE* and its common *APOE* genotypes on the differential associations of Alzheimer’s dementia, the progressively harmful or protective molecular mechanisms that may account for these differences, and the importance of discovering interventions to safely and sufficiently target those factors to the treatment and prevention of AD. As we predicted from the likely inclusion of neuropathologically misclassified cases and controls in the clinical group, APOE3/4 and 4/4 ORs were significantly lower than those in the neuropathologically confirmed autopsy group and roughly comparable to those from numerous case-control studies in which neuropathological (or biomarker) measurements were not required to confirm the presence or absence of AD.^1^ APOE2/4, 3/4, and 4/4 Alzheimer’s dementia ORs relative to APOE3/3 were 2.47, 3.55, and 10.70, in the neuropathologically unconfirmed group (***Table 1***), and 2.47, 3.78, and 12.02 in the combined group (***Supplementary Table 4***), respectively.

#### Impact of APOE4 allelic dose

As shown in ***Table 1 and Supplementary Tables 3 and 4***, APOE4 allelic dose in APOE2 non-carriers was associated with significantly greater ORs in the neuropathologically confirmed autopsy, neuropathologically unconfirmed clinical, and combined groups. Using an additive genetic model, ORs in the neuropathologically confirmed (6.00 OR [95% CI=5.06-7.12] P=3X10^−90^), unconfirmed (3.43 OR [95% CI=3.26-3.60] P<1X10^−300^), and combined group (3.61 OR [95% CI=3.37-3.87] P=2X10^−290^) were similar to corresponding APOE3/4 OR relative to APOE3/3. As shown in ***Supplementary Tables 3 and 4***, findings were not significantly different after adjustment for age or sex in the neuropathologically confirmed or unconfirmed group or after adjustment for age, sex, or neuropathologically confirmed autopsy/unconfirmed non-autopsy group. Using the resulting beta estimates, the 36.0 OR in the neuropathologically confirmed APOE4 homozygotes was similar to the 31.2 OR shown in Table 1.

#### Impact of neuropathologically confirmed autopsy versus unconfirmed non-group on Ors

We originally postulated that APOE2 and APOE4 allelic dose would have a more profound impact on Alzheimer’s dementia ORs in neuropathologically confirmed than unconfirmed cases and controls due to the exclusion of clinically diagnosed cases who did not meet neuropathological criteria for AD and unimpaired controls who met criteria for (preclinical) AD, which could dilute OR estimates in the unconfirmed group; and to known differences between APOE4 carriers and non-carriers in the neuropathological classification of cases (more likely in APOE4 non-carriers) and controls (more likely in APOE4 carriers), which could lead to systematically underestimation ORs in the unconfirmed group. Since the ADGC does not have information from most of the neuropathologically misclassified cases and controls, it is not possible to clarify whether the more profound impact of different APOE genotypes and allelic doses on Alzheimer’s dementia ORs is solely attributable to the exclusion of neuropathologically misclassified cases and controls or also on participation in a brain donation program. As previously noted, ORs for each APOE genotype and allelic dose were not significantly different after adjustment for neuropathologically confirmed autopsy/unconfirmed non-autopsy group.

#### Impact of APOE2 and APOE4 allelic dose on the classification of cases and controls

A sensitivity analysis was performed to clarify the impact of age, sex, and neuropathologically confirmed autopsy/unconfirmed autopsy group on the accuracy to classify cases and controls. The Receiver Operating Characteristic (ROC) Curves in ***Supplementary Figure 1*** show the impact of APOE2 and APOE4 allelic doses on the classification of cases and controls in the neuropathologically confirmed autopsy and unconfirmed non-autopsy groups. APOE2 and APOE4 allelic doses were each associated with significantly greater Areas Under the Curve (AUC), an indicator of classification accuracy, in the neuropathologically confirmed and unconfirmed groups, and APOE4 allelic dose had a greater impact than APOE2 allelic dose on AUCs in both groups, as reflected by the variance importance scores, below. APOE2 allelic dose was associated with a significantly greater AUC in the neuropathologically confirmed autopsy than unconfirmed non-autopsy group (AUC 0.68 [95%CI 0.65-70 versus 0.58 [95%CI 0.50-0.65]), corresponding to variance importance scores of 24.0 versus 9.0, respectively. AUCs for APOE4 allelic dose were not significantly different in the neuropathologically confirmed autopsy and non-autopsy groups, as reflected by AUC 0.77 (95%CI 0.75-79) versus 0.75 (95%CI 0.74-0.77) and corresponding variance importance scores of 43.9 and 32.3.

### Impact of *APOE* genotypes and allelic doses on four other neuropathological disease ORs

Other neuropathological diseases are relatively common in older adults with and without Alzheimer’s dementia. ***Supplementary Table 5*** shows the number of persons in the neuropathologically confirmed case-control group with and without four commonly assessed neuropathological diagnoses, including congophilic amyloid angiopathy (CAA), Lewy Body Disease (LBD), Vascular Brain Injury (VBI), and hippocampal sclerosis (HS). (Since TDP-43 pathology and microinfarcts were not characterized in many of the participants, they were not included in our analysis.) CAA, LBD, VBI, and HS were present in 94, 88, 78, and 83% of the Alzheimer’s dementia cases and 70, 22, 12, and 14% of the unimpaired non-AD controls. ***Supplementary Tables 6 and 7*** show CAA, LBD, VBI, and HS ORs for each APOE genotype, compared to APOE3/3, and for allelic dose, before and after adjustment for age, sex, and the neuropathological diagnosis of AD. APOE2 allelic dose was not associated with a significantly lower OR for any of these diseases, before or after adjustment for presence or absence of AD. While APOE4 allelic dose was not significantly associated with significantly higher VBI and HS ORs, it was associated with significantly higher CAA and LBD ORs, before and after adjustment for age, sex, and presence or absence of AD.

### Impact of APOE genotypes on ages at dementia onset

The estimated mean ages at Alzheimer’s dementia onset for each genotype shown in **Supplementary Table 2** are consistent with previously reported findings. In neuropathologically confirmed Alzheimer’s dementia cases with available onset ages, APOE4/4, 3/4, 2/4, 3/3, and combined 2/3 and 2/2 genotypes were associated with progressively older ages at Alzheimer’s dementia onset, ranging from 69.9 ± 6.1 years in the APOE4/4 genotype to 79.3±9.0 years in the combined APOE2/3 and 2/2 group. In the neuropathologically unconfirmed and combined cases, APOE4/4, 3/4, 2/4, 3/3, and combined 2/3 and 2/2 genotypes were also associated with progressively older ages at Alzheimer’s dementia onset, ranging from 69.5±5.9 years in the combined APOE4/4 homozygote group to 77.7±8.5 years in the combined APOE2/3 and 2/2 group. The Kaplan-Meier curves in ***Figure 1*** show the percentage of persons in the neuropathologically confirmed case-control group with each APOE genotype, including APOE2/2, who remained free from Alzheimer’s dementia at different ages. While there was some overlap between the 95% CIs in the APOE2/2 and 2/3 plots due to the small size of and relatively large CI for the APOE2/2 group, the Kaplan-Meier plots in ***Figure 1*** confirmed a relationship between APOE2 allelic dose and freedom from Alzheimer’s dementia survival at older ages.

**Figure 1.**
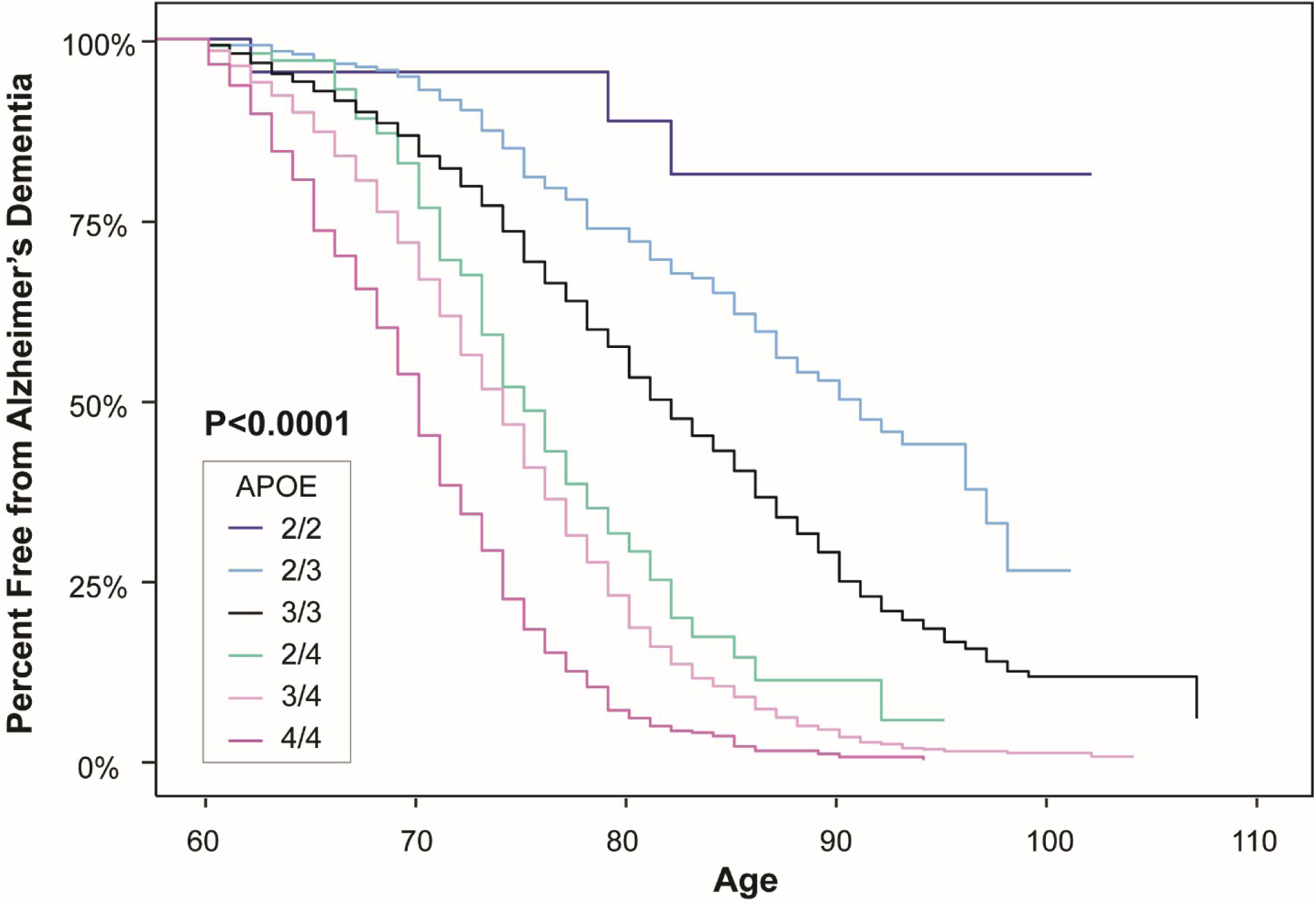
The Percentage of Persons with Each APOE Genotype in the Neuropathologically Confirmed Group Who Remained Free from Alzheimer’s Dementia as a Function of Age. Kaplan-Meier curves were generated from Alzheimer’s dementia cases and cognitively unimpaired non-AD controls in the neuropathologically confirmed group.

### Impact of *APOE* genotypes on neuritic plaque and neurofibrillary tangle severity

***Supplementary Table 8 and Supplementary Figure 2***, provide information about CERAD (neuritic Aβ plaque) scores and Braak (tau tangle) stage in the neuropathologically verified case-control group. APOE genotype (APOE2/2<2/3<3/3<2/4<3/4<4/4) (and consequently allelic dose) were associated with greater Aβ plaque and tau tangle severity, before and after adjustment for age at death and sex. Our findings could be attributable to the proportion of cases to controls in each genetic group, their direct or indirect impact on these neuropathological features, potentially confounding differential effects of AD onset and duration on Braak stage, or a combination of these and other factors. While the difference between CERAD scores in the small APOE2/2 and 2/3 groups were not significant (1.05±1.10 versus 1.33±1.31, P=0.35), APOE2 homozygotes were distinguished from the APOE2/3 group by a significantly lower Braak stage (2.26±1.63 versus 3.16±1.69, P<0.05), before and after adjustment for age and sex. Compared to the APOE3/3 group, the APOE2/2 and APOE2/3 groups had significantly lower CERAD scores and Braak stages, and the APOE2/4, 3/4, and 4/4 genotypes had significantly higher CERAD scores and Braak stages, before and after adjustment for age at death and sex.

Residual effects of each APOE genotype on Braak stage, relative to APOE2/2, 3/3, and 4/4, after controlling CERAD scores are depicted in terms of beta (linear regression) coefficients in ***Table 3***. Relatively protective or harmful effects are reflected by negative or positive beta coefficients, respectively. Compared to APOE2/3 or APOE3/3, only the APOE3/4 and 4/4 genotypes demonstrated progressively harmful residual effects on Braak stage. Compared to APOE4/4, the APOE2/2, 2/3, 3/3, 2/4, and 3/4 genotypes had significant and progressively protective residual effects on Braak stage with beta coefficients of −1.32, −0.43, −0.25, −0.26, and −0.10, respectively (P<0.05), such that the protective effect was more than three times greater in the APOE2/2 than APOE2/3 group. This finding supports the possibility that APOE variants have differential effects on tau tangle severity, even after controlling for neuritic Aβ plaque severity.

## DISCUSSION

This study demonstrates an exceptionally low likelihood of Alzheimer’s dementia in APOE2 homozygotes in a large group of clinically and neuropathologically characterized cases and controls. The large number of cases and controls in the neuropathologically unconfirmed and combined groups enabled us to demonstrate a significantly greater impact of APOE2/2, 2/3, 3/3, 3/4, and 4/4 genotypes on Alzheimer’s dementia ORs in the neuropathologically confirmed group—and to suggest that the greater impact may be attributable to the exclusion of cases without significant AD neuropathology, as well as the exclusion of controls with preclinical AD neuropathology.^11^ This study provides updated ORs for Alzheimer’s dementia for each of the six common APOE genotypes, APOE2 allelic dose, and APOE4 allelic dose on, on the differential risk of Alzheimer’s dementia., free from the confounding effects of clinically misdiagnosed cases and controls, and it demonstrates an additional impact of APOE4 but not APOE2 allelic dose on two other neuropathological disease (CAA and DLB) ORs. The study supports known effects of *APOE* genotypes on standard measures of neuritic Aβ plaque and tau tangle severity^12^ and suggests progressively protective residual effects on Braak stage in APOE3/4, 2/4, 3/3, 2/3, and 2/2 groups compared to APOE4/4 homozygotes.

The APOE2/2 genotype was associated with a significantly lower 0.34 Alzheimer’s dementia OR compared to the relatively low odds APOE2/3 genotype (95% CI 0.12-0.95), an extremely low 0.13 OR compared to the most common APOE3/3 genotype (95% CI 0.05-0.36), and an exceptionally low 0.004 OR compared to the highest odds APOE4/4 genotype (95% CI 0.001-0.014) in those neuropathologically confirmed subjects. In other words, persons with the APOE2/2 genotype had a 66% lower OR than those with the APOE2/3 genotype, 87% lower than those with the APOE3/3 genotype, and 99.6% (95% CI 98.6-99.9%) lower than those with APOE4/4 group. These findings highlight the impact of *APOE* and its variants on the risk of AD and the potential impact of *APOE*-modifying interventions on its treatment and prevention.

The APOE2/2 genotype has recently been suggested to be associated with more severe pathology in primary tauopathies, including progressive supranuclear palsy, corticobasal degeneration, and a mouse model of human tau over-expression.^13^ On the other hand, APOE2/2 and APOE3/3ch genotypes could be associated with less severe tau pathology in AD (a secondary tauopathy) due to a direct or indirect (e.g., amyloid and neuroinflammation-mediated) effects, as noted above. While the current study did not assess the impact of APOE2 allelic dose on ORs for these primary tauopathies, it found no association between APOE2 allele dose and ORs for four other disease (CAA, DLB, VBI, and HS). In contrast, APOE4 allelic dose was associated with significantly higher ORs for CAA and DLB, before and after adjustment for the presence or absence of AD, and not with VBI or HS.

As previously noted, we and our colleagues recently found as association between two copies of the rare APOE3ch mutation and resistance to the clinical onset of AD in an Aβ_42_-overproducing PSEN1 E280A mutation carrier from the world’s largest autosomal dominant AD kindred.^5^ Interestingly, this individual had unusually high PET measurements of Aβ plaque burden, relatively limited PET measurements of paired helical filament tau (neurofibrillary tangle) burden, minimal cerebral glucose hypometabolism in AD-affected brain regions, and like some APOE2 homozygotes, Type III hyperlipoproteinemia.^5,6^ We postulate that the contributions of *APOE* and its variants to the differential risk of Alzheimer’s dementia are not solely attributable to their impact on the density of Aβ plaques but also to their direct or indirect impact on downstream pathogenic events. In experimental studies, the ApoEch protein was associated with reduced Aβ_42_ aggregation and a differential impact of ApoE isoforms (ApoE4>3>2>>3ch) on heparin binding and demonstrated the ability of a targeted antibody to lower wildtype ApoE3 binding to heparin.^5^ Based on studies implicating heparin sulfate polyglycan (HSPG) on Aβ aggregation, Aβ-mediated microglial response, the neuronal uptake and potential propagation of tau,^14^ and neurodegeneration, we postulated that the ApoE binding to HSPG could have potential roles in the pathogenesis, treatment, and prevention of AD.^5^

Together our APOE2/2 and APOE3ch/3ch studies underscore the need to clarify and target the factors by which APOE and its variants account for this differential risk, treatment and prevention of AD. It remains to be shown whether the differential effects of *APOE* variants on the risk of Alzheimer’s dementia are related to their recognized effects on Aβ oligomerization, morphology, or clearance, their suggested effects on *TREM2*-mediated microglial response or tau pathology, low density lipoprotein receptor (LDLR) binding, HSPG binding, mitochondrial function, neurodegeneration, or a combination of these and other effects.^5,7,15-20^ It also remains to be shown whether it would help to increase or, as some of us suggest, decrease ApoE expression in brain.

*APOE*, its variants, and the right *APOE*-modifying treatments could influence Aβ oligomerization or clearance, the morphology of Aβ aggregates, *TREM2*-mediated neuroinflammation, development or spread of tau pathology, mitochondrial function, neurodegeneration, or a combination of these or other processes. For instance, like their loss-of-function effects on LDLR binding, heparin sulfate polyglutamate (HSPG) binding, and Aβ oligomerization,^5-7^ a non-additive protective effect of APOE2 in reducing the expression of a microglial aging signature (HuMi Aged geneset),^21^ APOE2 variants might be associated with a relative loss in one or more functions that are critically involved in the development of AD. If so, *APOE* and its variants could differ in the extent of their pathogenic functions (APOE4/4>3/4>2/4>3/3>2/3>2/2 and APOEch/ch>no APOE). We postulate that genetic, drug or immune treatments that safely and sufficiently inhibit the expression of *APOE* or its relevant functions in brain might have a significant impact on the treatment and prevention of AD. Evidence from a person without *APOE* function due to homozygosity for an ablative *APOE* frameshift mutation^22^ and the availability of dyslipidemia treatments support the potential tolerability of this approach. Additional research is needed to clarify the mechanisms by which homozygosity for APOE2 and APOEch are associated with an exceptionally low risk of AD dementia, Gene editing, protein-reducing, protein-modifying, or other treatments that safely and sufficiently replicate protective effects of the APOE2/2 genotypes could help to prevent the clinical onset of AD.

The study has several limitations. First, while the unusually large number of participants in the overall study, there was a relatively small number of APOE2 homozygotes. Despite the resulting limitation in statistical power, it did not prevent us from demonstrating significant effects in association with the APOE2/2 genotype and APOE2 allelic dose. Second, as previously noted, since we do not have information from most of the participating brain banks’ neuropathologically misclassified cases and controls, we are not able to clarify with the more profound impact of different APOE genotypes and allelic doses on Alzheimer’s dementia ORs is solely attributable to the exclusion of neuropathologically misclassified cases and controls or also to any ascertain biases related to participation in a brain donation program. However, we were able to quantify and distinguish the impact of APOE2 versus APOE4 allelic doses on the classification of cases and controls in both the neuropathologically confirmed and unconfirmed groups.) Third, we cannot exclude an impact of differential disease onset, duration, or survival on Alzheimer’s dementia ORs and measures of AD pathology. Fourth, findings from our Non-Hispanic White groups cannot yet be extended to other ethnic and racial groups.^4^

Prospective cohort studies that include persons irrespective of their cognitive stage or neuropathological diagnosis are needed to clarify the absolute lifetime risk of neuropathologically confirmed Alzheimer’s dementia for each *APOE* genotype. Similarly, neuropathological studies that include brain donors irrespective of their cognitive stage or neuropathological diagnosis are needed to further clarify the impact of APOE2 gene dose on tau pathology and neurodegeneration, including the extent to which this impact is or is not mediated through its effect on Aβ pathology.^8,23^ Since the reported prevalence and impact of different APOE genotypes on Alzheimer’s dementia risk depends in part on age, race, ethnicity, geographic location, education, and dementia severity, these estimates are likely to vary in different populations. Based on our selection criteria, this study does not provide information about the percentage of *APOE* genotypes in cognitively unimpaired persons with neuropathological or biomarker evidence of preclinical AD, the percentage of persons who met criteria for MCI with or without neuropathological or biomarker evidence of AD, or the percentage of persons with a primary diagnosis of other neurodegenerative disorders. The impact of different APOE genotypes on estimated ages at Alzheimer’s dementia onset may have been greater if standardized prospective assessments had been used to estimate onset ages at every site, if the study included more research participants who developed Alzheimer’s dementia at younger ages (e.g., preferentially reducing onset ages in the APOE4 carrier groups), and if it included more participants who developed Alzheimer’s dementia at the oldest ages, e.g., preferentially increasing ages onset ages in the APOE4 non-carrier groups.

This study supports the complementary value of risk factor assessments in large neuropathologically confirmed autopsy and even larger neuropathologically unconfirmed clinical groups. In a previous autopsy study, we found that 25% of persons with the clinical diagnosis of mild-to-moderate Alzheimer’s dementia lacked at least moderately frequent neuritic plaques, one of the cardinal features of AD, including 37% of APOE4 non-carriers and 13% of carriers, findings that are consistent with those in living patients.^9,10^ In a meta-analysis of brain imaging studies, about 25% of cognitively unimpaired older adults have brain imaging evidence consistent with at least moderately frequent plaques. By investigating clinically characterized cases who are confirmed to have AD to unimpaired controls who are confirmed to be free of AD, it may be possible to address potentially confounding effects of the risk factor on the presence or absence of AD and underscore the potential impact of an intervention to prevent both AD and its clinical consequences. By investigating an even larger number of cases and controls without biological confirmation, it may be possible to clarify risk factors without potentially confounding effects on the underlying disease with improved statistical power and greater generalizability to understudied populations. Brain imaging and fluid biomarkers have the potential increase the size and generalizability of findings in case-control studies of AD.

Additional research is needed to clarify the mechanism by which *APOE*, and its variants contribute to the pathogenesis and potential treatment and prevention of AD. There is a critical need to discover treatments that account for impact of genotypes on the differential risk of Alzheimer’s dementia, including those that may account for a profound resistance to Alzheimer’s dementia in APOE2 and APOEch homozygotes, and to establish their value in the treatment and prevention of AD.

In conclusion, homozygosity for the APOE2 allele appears to be associated with an exceptionally low likelihood of AD dementia, *APOE* genotypes have an extraordinary impact on Alzheimer’s dementia ORs, and treatments that target *APOE* and its variants could have an extraordinary impact on the treatment and prevention of this terrible disease.

## METHODS

### Subjects

Our primary analysis capitalized on data from 5,007 brain donors in the ADGC’s “neuropathologically confirmed autopsy group” including 4,018 cases who met clinical and neuropathological criteria for Alzheimer’s dementia and 989 cognitively unimpaired controls who did not meet neuropathological criteria for AD; 5 of the cases and 19 of the controls had the APOE2/2 genotype. There were 283 brain donors in the ADGC’s “neuropathologically misclassified autopsy group” including 123 cases who met clinical criteria for probable Alzheimer’s dementia but not meet neuropathological criteria for AD and 160 unimpaired controls who met neuropathological criteria for AD. The entire autopsy group consisted of unrelated cases and controls. The ADGC’s “neuropathologically unconfirmed clinical group” contained 23,857 living research participants including 10,430 cases who met clinical criteria for probable Alzheimer’s dementia and 13,427 cognitively unimpaired controls; and the combined (overall autopsy and clinical subjects) group contained 29,147 research participants including 14,571 cases and 14,576 controls. Data from the neuropathologically confirmed autopsy group were used in our primary analyses; data from the other groups were in post hoc comparisons with that autopsy group. Due to the prioritized ascertainment of those cases who met clinical and neuropathological criteria for Alzheimer’s dementia and those unimpaired controls without AD, the number of misclassified cases and controls available through the AD Genetics Consortium^12^ is much smaller than our estimated number of neuropathologically misclassified cases and controls,^8-11^ limiting our ability to clarify the impact of brain donation on APOE ORs in our post-hoc analyses. The number of cases and controls for each APOE genotype in the ADGC’s neuropathologically confirmed autopsy group, neuropathologically misclassified autopsy group, neuropathologically unconfirmed clinical group, and combined autopsy and clinical group is shown in ***Supplementary Table 1***.

### Phenotypic Evaluation

Brain samples, extracted DNA, and demographic, clinical and neuropathological data from clinically and neuropathologically characterized brain donors were assembled by the AD Genetics Consortium (ADGC) in conjunction with past and present National Institute on Aging (NIA)-sponsored AD Centers, Banner Sun Health Research Institute and the Translational Genomics Research Institute (TGen), the Adult Changes in Thought Study (ACT), the University of Miami’s Brain Endowment Bank and Hussman Institute for Human Genomics, the Late-onset Alzheimer’s Disease Family Study, the Religious Orders Study and Memory Aging Project (ROS-MAP), the Vanderbilt University Center for Human Genetics Research, and the National Alzheimer’s Coordinating Center (NACC); *APOE* genotypes were characterized at the National Cellular Repository for AD (NCRAD) or TGen. Neuritic plaque burden was scored using the Consortium to Establish a Registry for AD (CERAD) 0-3 point (none, sparse, moderate, or frequent plaque) rating system, the spatial extent of neurofibrillary tangle (paired helical filament [PHF] tau) burden was scored using the Braak 0-VI staging system, and neuropathological data were reviewed and harmonized by a single neuropathologist (Thomas Montine).^12^ Neuropathological data in the autopsy group was collected at each site according to consensus guidelines at the time of brain autopsy. Cases with neuropathologic evidence of disease other than AD neuropathologic change, with or without common co-morbid lesions, were excluded. CERAD score, which provide an indicator of neuritic (Aβ) plaque severity, and Braak stage, which provides an indicator of neurofibrillary (tau) tangle spatial extent and severity, were available in every participant in the ADGC’s neuropathologically confirmed case-control autopsy group. Data regarding the presence or absence of four other neuropathological diagnoses commonly found in persons with AD, including congophilic amyloid angiopathy (CAA), Lewy body disease (LBD), vascular brain injury (VBI), and hippocampal sclerosis (HS) was most of the participants. Since TDP-43 proteinopathy and microinfarcts were not available in most of the brain donors (many of whom came to autopsy prior to development of TDP-43 proteinopathy), these neuropathological diseases were not included in our analysis. CAA, LBD, VBI, and HS were present in 84, 43, 38, and 21% of the brain donors, including Alzheimer’s dementia cases and unimpaired non-AD controls.

The Alzheimer’s dementia cases met DSM-IV or NINCDS/ADRDA criteria for dementia^24^ and, when available, had Clinical Diagnostic Ratings (CDRs) greater than zero before they died;^25^ they either met NIA/Reagan neuropathological criteria for intermediate-to-high likelihood AD or had both moderate-to-frequent neuritic plaque scores (i.e., CERAD score 2-3) and spatially extensive neurofibrillary tangle burden (i.e., Braak stage III-VI).^26-28^ The controls did not meet clinical criteria for dementia or mild cognitive impairment (MCI) and, when available, had a CDR of zero, within two years before they died; they met neuropathological criteria for low-likelihood AD, had sparse neuritic plaques (CERAD score 1) and spatially limited tangle burden (Braak Stages 0-II), or had no neuritic plaques (CERAD score 0) and no more than moderately extensive tangle burden (Braak stages 0-IV).

For comparative purposes, we subsequently analyzed *APOE* and other relevant data in clinically diagnosed but neuropathologically unexamined participants assembled by the ADGC from non-Hispanic whites. Their clinical, neuropathological, and demographic assessment procedures and characteristics were detailed in a previous report.^29^ Ages at clinical onset (when available) and last clinical examination are shown for each *APOE* genotype in ***Supplementary Tables 2 and 3***.

### Statistical Analysis

For the assessment of ORs associated with each *APOE* genotype, we coded the *APOE* genotype of interest as 1, coded the reference genotype as 0 (APOE2/2, APOE3/3, or APOE4/4), and treated other genotypes as missing. We used clinical diagnosis as a binary outcome for the neuropathologically confirmed, clinical, and combined group. Alzheimer’s dementia Odds Ratios (ORs) and 95% confidence intervals (95% CIs) for each *APOE* genotype were computed compared to a reference *APOE* genotype. We conducted genotypic association tests for each *APOE* genotype compared to a reference genotype or allelic association tests for the APOE2 or the APOE4 allele using the binary outcome with and without covariate adjustment for age and sex. For neuropathologically confirmed and misclassified autopsy groups, we computed ORs under a logistic regression using a generalized linear model (GLM). For clinical and combined groups, we further accounted for family structure due to containing families from the Multi-Institutional Research on Alzheimer’s Genetic Epidemiology (MIRAGE) and National Institute on Aging Late-onset AD (NIA-LOAD) study^30^ under a logistic regression using generalized estimating equations (GEE). For the assessment of APOE2 allelic dose, we coded APOE3/3, APOE2/3, and APOE2/2 as 0, 1, 2, respectively in the APOE4 non-carriers; and for the assessment of APOE4 allelic dose, we coded APOE3/3, APOE3/4, and APOE4/4 as 0, 1, 2, respectively, in the APOE2 non-carriers. We conducted allelic association tests for the APOE2 and APOE4 under a logistic regression using a GLM for AD, LBD, VBI, HS and CAA. We used a sensitivity analysis, receiver operating characteristics (ROC) curves, and areas under the curve (AUCs) to characterize and compare the contribution of APOE2 and APOE4 allelic doses on the classification of cases and controls in the neuropathologically confirmed and unconfirmed groups. To rank predictors based on their contributions to the logistic regression model, we calculated variance importance scores using the *varImp* option in R, which the importance score is ranged from 0 to 100%.

Data from the neuropathological cohort were used to generate the Kaplan-Meier curves for each APOE genotype that are shown in ***Figure 1***. The curves indicate the percentage of neuropathologically confirmed cases and controls who remained free from Alzheimer’s dementia as a function of age. When estimated ages at dementia onset were not available, ages at death were used as a proxy.

CERAD scores and Braak stages were quantified for each *APOE* genotype in the aggregate group of autopsied cases and controls from both neuropathologically confirmed and misclassified autopsy groups. Linear regression using the GLM model was used to assess the effect of each *APOE* genotype comparing to the reference genotype for CERAD and Braak measurements as quantitative outcomes, repeated using CERAD scores as a covariate to assess residual effects of Braak stage.

## Data Availability

N/A

## ACKNOWLEDGEMENTS

This study was supported by the National Institute on Aging (NIA) grant grants RF1AG057519, P30-AG10161, R01AG15819, R01AG17917, P30 AG010133 and P30-AG19610 for the Arizona Alzheimer’s Disease Core Center; Arizona Department of Health Services contract 211002 for the Arizona Alzheimer’s Research Center; Arizona Biomedical Research Commission contracts 4001 0011, 05-901, and 1001 to the Arizona Parkinson’s Disease Consortium; and the Michael J. Fox Foundation for Parkinson’s Research. The Alzheimer’s Disease Genetics Consortium supported the collection of samples used in this study through NIA U01-AG032984 and RC2AG036528. Data for this study were prepared, archived, and distributed by the National Institute on Aging Alzheimer’s Disease Data Storage Site (NIAGADS) at the University of Pennsylvania and funded by NIA grant U24-AG041689. This work was also supported by NIA grants R01-AG048927, R01-AG032990, RF-AG051504, U01-AG046139, AG030653, AG041718, AG005133,

P50 AG005136 (UW ADRC NP Core) and U01 AG006781 (Adult Changes in Thought Study) and the Nancy and Buster Alvord Endowment (CDK), as well as by National Institute of Neurological Disorders and Stroke (NINDS) grant R01-NS080820 (to NET). The Brain and Body Donation Program at Sun Health Research Institute (Sun City, AZ, USA) is supported by NINDS grant U24-NS072026 for the National Brain and Tissue Resource for Parkinson’s Disease and Related disorders. National Institute of Neurological Disorders and Stroke NINDS grants (UG3NS115605 and RF1NS110048).

## Notes

### Competing Interest Statement

The authors have declared no competing interest.

## REFERENCES

1. Coon, K.D., et al.A high-density whole-genome association study reveals that APOE is the major susceptibility gene for sporadic late-onset Alzheimer’s disease. J Clin Psychiatry 68, 613–618 (2007).

2. Corder, E.H., et al. Protective effect of apolipoprotein E type 2 allele for late onset Alzheimer disease. Nat Genet 7, 180–184 (1994).

3. Corder, E.H., et al. Gene dose of apolipoprotein E type 4 allele and the risk of Alzheimer’s disease in late onset families. Science 261, 921–923 (1993).

4. Farrer, L.A., et al. Effects of age, sex, and ethnicity on the association between apolipoprotein E genotype and Alzheimer disease. A meta-analysis. APOE and Alzheimer Disease Meta Analysis Consortium. JAMA 278, 1349–1356 (1997).

5. Arboleda-Velasquez, J.F., et al. Resistance to autosomal dominant Alzheimer’s in an APOE3-Christchurch homozygote. (submitted).

6. Mahley, R.W., Huang, Y. & Rall, S.C., Jr. Pathogenesis of type III hyperlipoproteinemia (dysbetalipoproteinemia). Questions, quandaries, and paradoxes. J Lipid Res 40, 1933–1949 (1999).

7. Hashimoto, T., et al.Apolipoprotein E, especially apolipoprotein E4, increases the oligomerization of amyloid beta peptide. J Neurosci 32, 15181–15192 (2012).

8. Monsell, S.E., et al. Characterizing Apolipoprotein E epsilon4 Carriers and Noncarriers Withthe Clinical Diagnosis of Mild to Moderate Alzheimer Dementia and Minimal beta-Amyloid Peptide Plaques. JAMA Neurol 72, 1124–1131 (2015).

9. Reiman, E.M., et al. Fibrillar amyloid-beta burden in cognitively normal people at 3 levels of genetic risk for Alzheimer’s disease. Proc Natl Acad Sci U S A 106, 6820–6825 (2009).

10. Jansen, W.J., et al. Prevalence of cerebral amyloid pathology in persons without dementia: a metaanalysis. JAMA 313, 1924–1938 (2015).

11. Escott-Price, V., Baker, E. & Shoai, M. Genetic analysis suggests high misassignment rates inclinical Alzheimer’s cases and controls. Neurobiology of Aging (in press).

12. Serrano-Pozo, A., Qian, J., Monsell, S.E., Betensky, R.A. & Hyman, B.T. APOEepsilon2 is associated with milder clinical and pathological Alzheimer disease. Ann Neurol 77, 917–929 (2015).

13. Zhao, N., et al. APOE epsilon2 is associated with increased tau pathology in primary tauopathy. Nat Commun 9, 4388 (2018).

14. Rauch, J.N., et al. Tau Internalization is Regulated by 6-O Sulfation on Heparan Sulfate Proteoglycans (HSPGs). Sci Rep 8, 6382 (2018).

15. Huynh, T.V., et al. Age-Dependent Effects of apoE Reduction Using Antisense Oligonucleotides in a Model of beta-amyloidosis. Neuron 96, 1013–1023 e1014 (2017).

16. Mahley, R.W. Apolipoprotein E: from cardiovascular disease to neurodegenerative disorders. J Mol Med (Berl) 94, 739–746 (2016).

17. Shi, Y. & Holtzman, D.M. Interplay between innate immunity and Alzheimer disease: APOE and TREM2 in the spotlight. Nat Rev Immunol 18, 759–772 (2018).

18. Shi, Y., et al. ApoE4 markedly exacerbates tau-mediated neurodegeneration in a mouse model of tauopathy. Nature 549, 523–527 (2017).

19. Wang, C., et al. Gain of toxic apolipoprotein E4 effects in human iPSC-derived neurons is ameliorated by a small-molecule structure corrector. Nat Med 24, 647–657 (2018).

20. Yamazaki, Y., Painter, M.M., Bu, G. & Kanekiyo, T. Apolipoprotein E as a Therapeutic Target in Alzheimer’s Disease: A Review of Basic Research and Clinical Evidence. CNS Drugs 30, 773–789 (2016).

21. Olah, M., et al. A transcriptomic atlas of aged human microglia. Nat Commun 9, 539 (2018).

22. Mak, A.C., et al. Effects of the absence of apolipoprotein e on lipoproteins, neurocognitive function, and retinal function. JAMA Neurol 71, 1228–1236 (2014).

23. Salloway, S. & Sperling, R. Understanding Conflicting Neuropathological Findings in Patients Clinically Diagnosed as Having Alzheimer Dementia. JAMA Neurol 72, 1106–1108 (2015).

24. Naj, A.C., et al. Common variants at MS4A4/MS4A6E, CD2AP, CD33 and EPHA1 are associated with late-onset Alzheimer’s disease. Nat Genet 43, 436–441 (2011).

25. Burke, W.J., et al. Reliability of the Washington University Clinical Dementia Rating. Arch Neurol 45, 31–32 (1988).

26. Mirra, S.S., Hart, M.N. & Terry, R.D. Making the diagnosis of Alzheimer’s disease. A primer for practicing pathologists. Arch Pathol Lab Med 117, 132–144 (1993).

27. Braak, H. & Braak, E. Neuropathological stageing of Alzheimer-related changes. Acta Neuropathol 82, 239–259 (1991).

28. Hyman, B.T. & Trojanowski, J.Q. Consensus recommendations for the postmortem diagnosis of Alzheimer disease from the National Institute on Aging and the Reagan Institute Working Group on diagnostic criteria for the neuropathological assessment of Alzheimer disease. J Neuropathol Exp Neurol 56, 1095–1097 (1997).

29. Kunkle, B.W., et al. Genetic meta-analysis of diagnosed Alzheimer’s disease identifies new risk loci and implicates Abeta, tau, immunity and lipid processing. Nat Genet 51, 414–430 (2019).

30. Jun, G., et al. Meta-analysis confirms CR1, CLU, and PICALM as alzheimer disease risk loci and reveals interactions with APOE genotypes. Arch Neurol 67, 1473–1484 (2010).

